# Mortality in Switzerland in 2021

**DOI:** 10.1101/2022.04.07.22273557

**Authors:** Isabella Locatelli, Valentin Rousson

## Abstract

**Objective:** To analyze mortality trends in Switzerland in 2021, the second year of the COVID-19 pandemic.

**Methods:** Using data from the Swiss Federal Statistical Office, we compared mortality in Switzerland in 2021 with that of previous years in terms of standardized weekly deaths, standardized (annual) mortality rates (overall and stratified by age and sex) and life expectancy.

**Results:** After a favorable first half of the year and a fairly standard second half in terms of mortality in Switzerland, the year 2021 ended with a wave of deaths of moderate intensity related to the 5th wave of COVID-19. Overall, and after a notable increase in mortality in 2020 (+9.2% compared to 2019), the pre-pandemic mortality level was approximately recovered in 2021 (+0.6% compared to 2019). Life expectancy, after declining by 10 months for men and 6 months for women in 2020, returned in 2021 to levels slightly above those of 2019 for women (85.7 years, +0.1 years from 2019) and regained 2018 levels for men (81.6 years, still -0.3 years from 2019). The age group responsible for the small remaining loss for men was the 50-70 age group, which had similar mortality in 2020 and 2021.

**Conclusions:** The second year of the COVID-19 pandemic in Switzerland was characterized by an approximate return to pre-pandemic mortality levels, with a faster recovery for women than for men compared to 2020.

## Introduction

The years 2020 and 2021 have seen the global spread of the COVID-19 pandemic caused by the severe acute respiratory syndrome coronavirus 2 (SARS-CoV-2). At the time of writing, nearly 500 million cases and more than 6 million deaths due to COVID-19 have been reported worldwide. In Switzerland, the pandemic has caused more than 13’000 deaths to date, representing 1’500 deaths per million inhabitants (https://www.worldometers.info/coronavirus/). A key indicator to assess the impact of the pandemic is all-cause mortality [1], a measure that does not suffer from over- or under-reporting and is able to capture both the direct and indirect effects of the pandemic on the population as a whole.

There is now a large literature studying the impact of the pandemic on all-cause mortality during the year 2020. The latter has been evaluated for a large number of countries both from the point of view of the excess mortality (or excess deaths) [1-4] and from the perspective of the loss of life expectancy [5-8]. For Switzerland, an excess mortality of 9.2% has been estimated in 2020, which was significant from the age of 70 for men and 75 for women. The loss of life expectancy was of 10 months for men and 6 months for women [9-10]. These results were consistent with those, now official, obtained a couple of months later by the Swiss Federal Statistical Office (<u>Espérance de vie | Office fédéral de la statistique (admin.ch)</u>) and with those of a large study on the loss of life expectancy in 29 countries [11].

On the other hand, while a few studies have been published concerning the excess death of (part of) the year 2021 [12-13], or globally for the period 2020-21 [14], no study has yet, to our knowledge, estimated the life expectancy achieved for the year 2021, nor analyzed the association between age and excess (or under-) mortality that year. The reason is certainly related to the time needed to obtain definitive data on deaths in a given country, and Switzerland is no exception, because part of the deaths are registered with delay by the Offices. We believe however that it is important to have early information on mortality during a pandemic, even if not with the highest precision, and consider that the impact of a second year of pandemic on mortality is a key issue for public health policy. This is why we have been following the weekly updates of deaths published by the Federal Statistical Office (FSO) since the beginning of 2022 and, after observing a certain convergence of the figures, we propose in this paper an initial estimate of mortality for 2021 on the basis of data that we consider to be sufficiently stable and almost definitive.

To quantify (excess) mortality in Switzerland in 2021, we use in this paper the indicators and methods that have already been used to analyze mortality in Switzerland in 2020 and in the first half of 2021 [8-10], i.e. standardized mortality rates, on an annual and weekly basis, and life expectancy. The values obtained for 2021 are compared with those for 2020 and 2019 by relative (rates) or absolute (life expectancy) differences. An analysis by sex and age group of the standardized rates is also provided in order to identify, as already done for 2020 [9], the age groups possibly more affected by this second year of the pandemic. Finally, an estimate of the excess deaths in each of the two years (and overall) is proposed for comparison with estimates obtained in the literature.

## Data

We used official data on deaths in Switzerland published by the Swiss FSO for 2000-2021. The annual number of deaths, separately for men and women, was available by 1-year age groups (with a last open class of 110+) until year 2020 (<u>Décès selon l’âge et le sexe, 1970-2020 - 1970-2020 | Tableau | Office fédéral de la statistique (admin.ch)</u>), while it was available by 5-years age groups (with a last open class of 90+) for each of the 52 weeks of 2021(<u>Décès selon la classe d’âge quinquennale, le sexe, la semaine et le canton - 4.1.2021-2.1.2022 | Tableau | Office fédéral de la statistique (admin.ch)</u>), and by 3 large age groups (0-64, 65-79, 80+) for the entire 2021 (January 1 - December 31) (<u>Décès | Office fédéral de la statistique (admin.ch)</u>). Weekly death data were also available for years 2015-2021 with a partition in five large age classes (0-19, 20-39, 40-64, 65-79, 80+) (<u>Décès par classe d’âge, semaine et canton - 29.12.2014-27.3.2022 | Tableau | Office fédéral de la statistique (admin.ch)</u>), last access April 5^th^ 2022. As mentioned in the Introduction, the death figures for 2021 were still provisional, as a number of deaths are recorded with a delay by the FSO. By convention, the year 2021 is divided into 52 weeks, the first of which begins on January 4, 2021 and the last of which ends on January 2, 2022. Therefore, we adjusted the weekly deaths to obtain the same totals for men and women as reported for annual deaths in each of the three age groups (0-64, 65-79, 80+). To facilitate comparisons across all years, we finally excluded 1/366 of deaths in the case of leap years (2000-’04-’08-’12-’16-’20).

The size of the Swiss population as of January 1, 2011-2021, stratified by sex and one-year age groups (with a last open class of 105+) was available in the FSO database (<u>Population résidante permanente selon l’âge, le sexe et la catégorie de nationalité, de 2010 à 2020 - 2010-2020 | Tableau | Office fédéral de la statistique (admin.ch)</u>). The size of the Swiss population as of January 1, 2000-2021, stratified by sex and one-year age groups (with a last open class of 110+), was also found in the Human Mortality Database (HMD) (<u>Human Mortality Database</u>). Since the difference for the overlapping years were negligible (mean relative difference of 0.007% with no difference exceeding 0.015%), we used HMD source for the years 2000-2010 and FSO source for the years 2011-2021. Life expectancy for both sexes was also available in the HMD database for the period 2000-2020 and was a useful gold standard for evaluating our method.

## Methods

Weekly mortality for the years 2015-2021 was analyzed using the Standardized Weekly Deaths (SWD), with the five available age classes (0-19, 20-30, 40-64, 65-79, 80+) and considering population as of January 1, 2021 as the reference, according to the method described in [10].

To compare mortality of 2021 with that of previous years we used (one-year of) age and sex (directly) Standardized Mortality Rates (SMR) [15-16], considering population as of January 1, 2021 as the reference. SMRs were calculated on the entire population and by sex and 10-year age groups (with a last open class of 90+). Mortality between two years was compared (globally and for a given sex/age group) using the relative change of SMR expressed in %. A complete description of SMR calculation, as well as of how to obtain a 95% confidence interval for a relative change between two SMRs can be found in [9].

Life expectancy (LE) at birth and at 65 years of age was obtained applying a piecewise exponential model [17] on death (and population) data stratified in 5-year age classes (with a last open class of 90+), i.e. using for all years the degree of age stratification available for 2021. The method is described in detail in [8]. For validation we compared LEs for years 2000-2020 obtained with our method and data with the ones published by the HMD. We found a mean relative difference of 0.05% with no difference exceeding 0.1%. Life expectancies between two years were then compared using absolute differences expressed in months.

## Results

Fig 1 shows the standardized weekly deaths (SWD) for the years 2015-2021 (reference: January 1, 2021). The SWD for a given week and year represents the number of deaths that would have occurred if the age-specific mortality rates observed that week and year were applied to the age structure of the 2021 population. This graph highlights the different waves of influenza-related deaths from 2015 to 2019, of which 2015 was the largest. In 2020, we clearly observe two waves of mortality attributable to COVID-19, where the second wave (in fall-winter 2020) was much higher and broader than the first (in spring 2020). In 2021, after the end of the second wave of COVID-19, we observed low mortality during the first half of the year, due to the absence of an influenza-related mortality wave and because the third and fourth waves of COVID-19 in Switzerland (spring and summer waves) had no visible impact on overall mortality. The second half of the year was characterized by a fairly standard mortality level with another moderate wave of mortality at the end of the year, corresponding to the 5th wave of COVID-19 in Switzerland.

**Figure 1:**
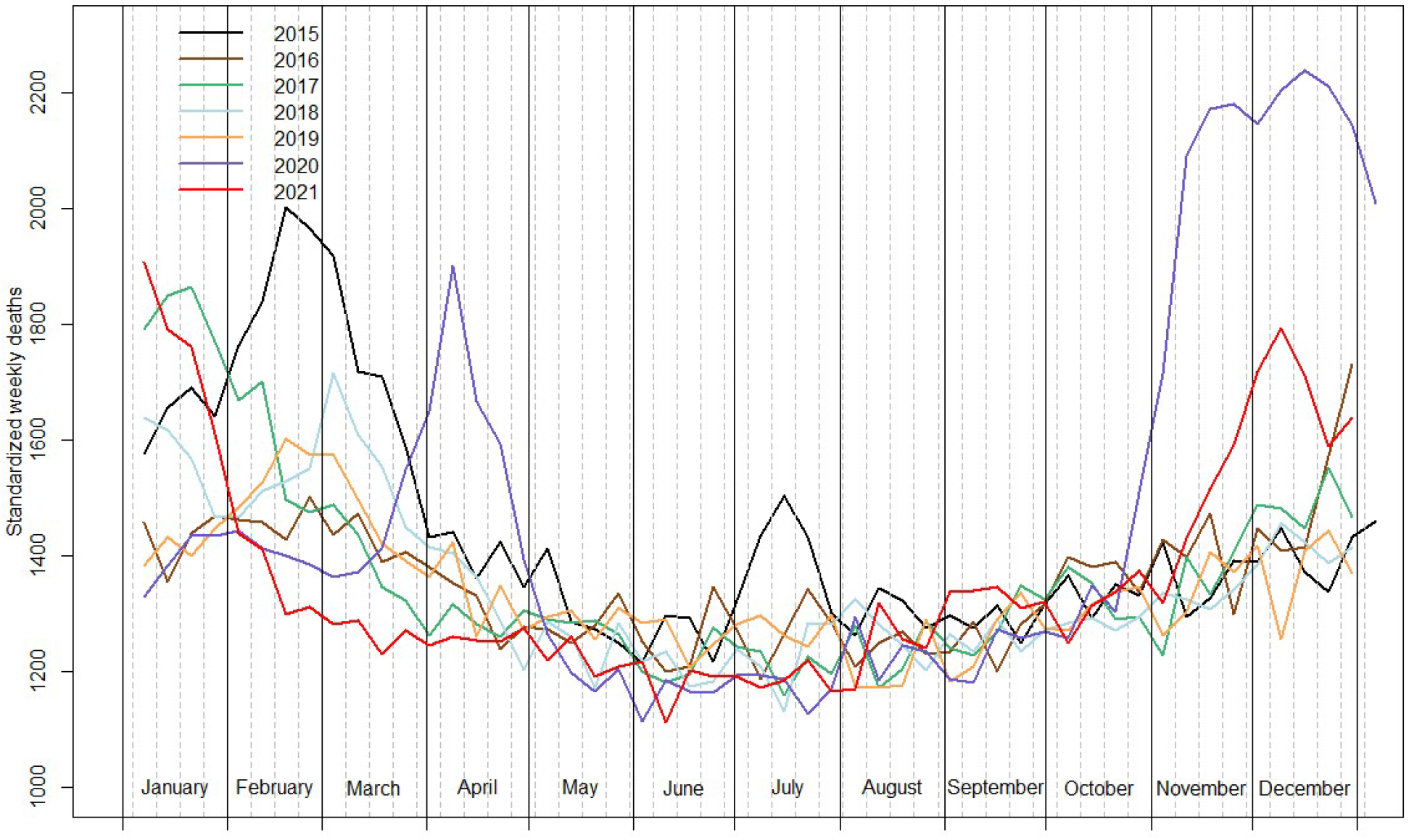
Standardized weekly deaths in Switzerland for the period 2015-2021, reference: January 1^st^ 2021 (data from the Swiss Federal Statistical Office)

The evolution of the number of deaths, crude mortality rates (CMR), and standardized mortality rates (SMR) (reference: January 1, 2021) between 2000 and 2021 is shown in Fig 2. We see a slightly increasing trend in the number of deaths and an approximately stable trend in the CMR between 2000 and 2019. The SMRs, on the other hand, have been decreasing steadily during these 20 years, indicating and illustrating a continuous progress regarding mortality. In 2020, all three indicators have increased. The absolute number of deaths and the CMR increased by 12.1% and 11.3%, respectively, in 2020, while SMR increased by 9.2%, 7.4% for women and 11.0% for men (Table 1), setting a return to the mortality level of the years 2014-15 [9]. This relative difference was largely the same, but in the opposite direction in 2021, bringing mortality back to 2018-19 levels (Fig 2, Table 1). However, whereas women have in 2021 (more than) fully recovered to 2019 mortality levels (SMR in 2021 of -0.5% compared to 2019), this was not quite the case for men (SMR in 2021 of +1.8% compared to 2019), who returned to 2018 levels. The excess deaths for 2020-21 was estimated by comparing the number of deaths in 2021 and 2020 to that in 2019, all three standardized to the population as of January 1, 2021. Using this method, the excess deaths for 2020-21 resulted in 6901 deaths, with 6473 for 2020 and 428 for 2021.

**Figure 2:**
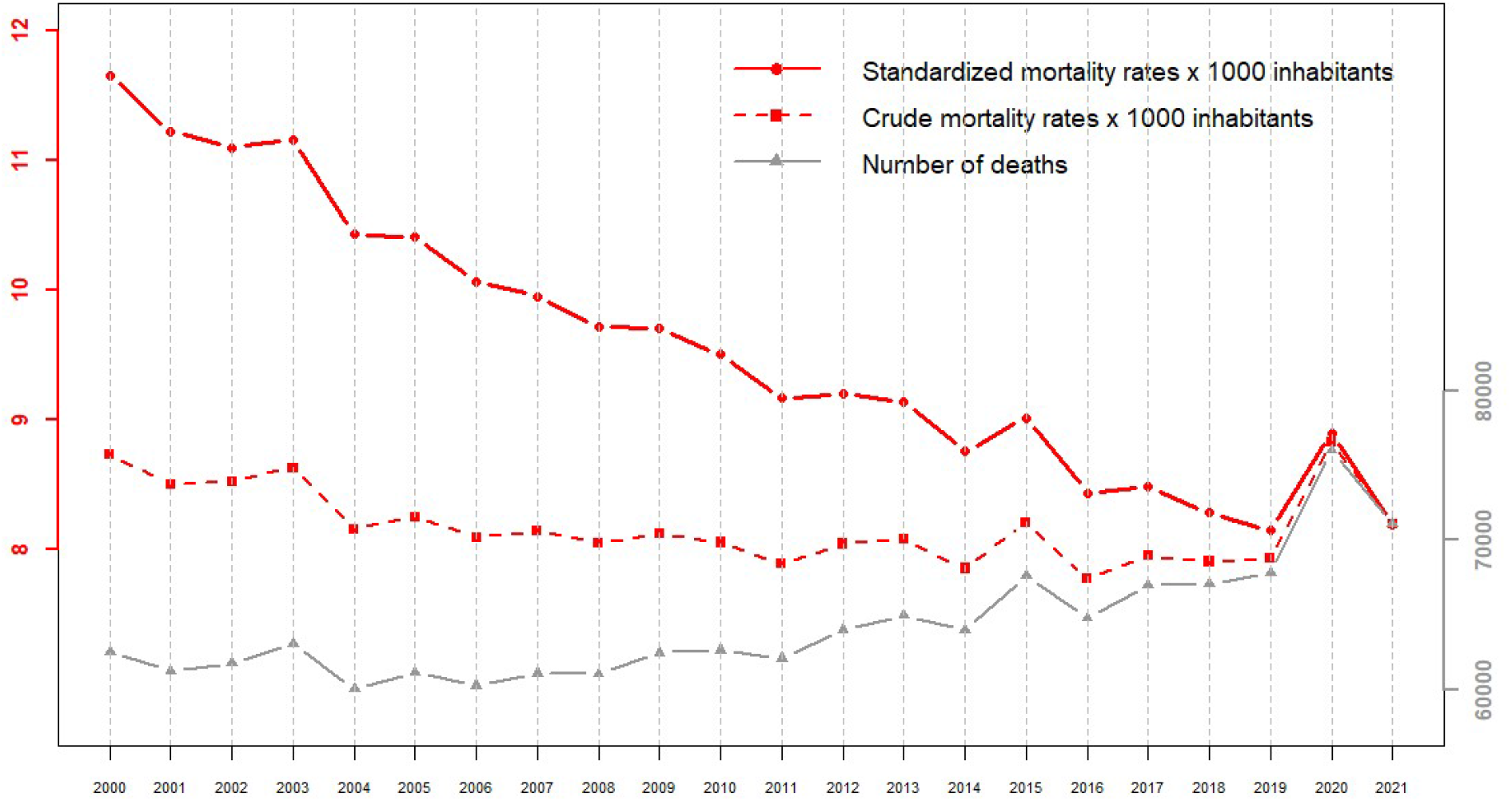
Number of deaths, crude mortality rates and standardized mortality rates (reference: January 1^st^ 2021) in Switzerland for the period 2000-2021 (data from the Swiss Federal Statistical Office and the Human Mortality Database)

**Table 1:** Summary of changes in standardized mortality rates (SMR) (%) and life expectancy (m=months) between 2019 and 2021

|  | SMR |  |  |  | Life Expectancy |  |  |  |  |  |
| --- | --- | --- | --- | --- | --- | --- | --- | --- | --- | --- |
|  | Total | Men | Women |  | Total |  | Men |  | Women |  |
|  |  |  |  |  | At birth | At 65 years | At birth | At 65 years | At birth | At 65 years |
|  |  |  |  | 2018 (years) | 83.5 | 21.2 | 81.6 | 19.8 | 85.4 | 22.6 |
|  |  |  |  | 2019 (years) | 83.7 | 21.3 | 81.9 | 19.9 | 85.6 | 22.6 |
|  |  |  |  | 2020 (years) | 83.1 | 20.7 | 81.0 | 19.1 | 85.1 | 22.2 |
|  |  |  |  | 2021 (years) | 83.6 | 21.2 | 81.6 | 19.8 | 85.7 | 22.7 |
| 2020/2019 (%) | +9.2 | +11.0 | +7.4 | 2020 - 2019 (m) | -7.9 | -7.2 | -10.1 | -8.8 | -5.8 | -5.7 |
| 2021/2020 (%) | -7.8 | -8.3 | -7.4 | 2021 - 2020 (m) | +6.7 | +6.7 | +7.2 | +7.4 | +6.2 | +6.0 |
| 2021/2019 (%) | +0.6 | +1.8 | -0.5 | 2021 - 2019 (m) | -1.2 | -0.6 | -2.9 | -1.4 | +0.5 | +0.3 |

Comparing the SMRs between 2019, 2020 and 2021 separately for both sexes and for 10-year age classes (last open class of 90+), we observe that the increased mortality in 2020 (compared to 2019) in age classes over 70 years [9] was almost perfectly counterbalanced by an equally large decrease in 2021 (compared to 2020). This led to statistically equivalent mortality in 2021 and 2019 in all age classes, with the exception of men aged 60-70, for whom mortality was higher in 2021 (Fig 3), while we also had a (non-significant) trend in this direction for men aged 50-60.

**Figure 3.**
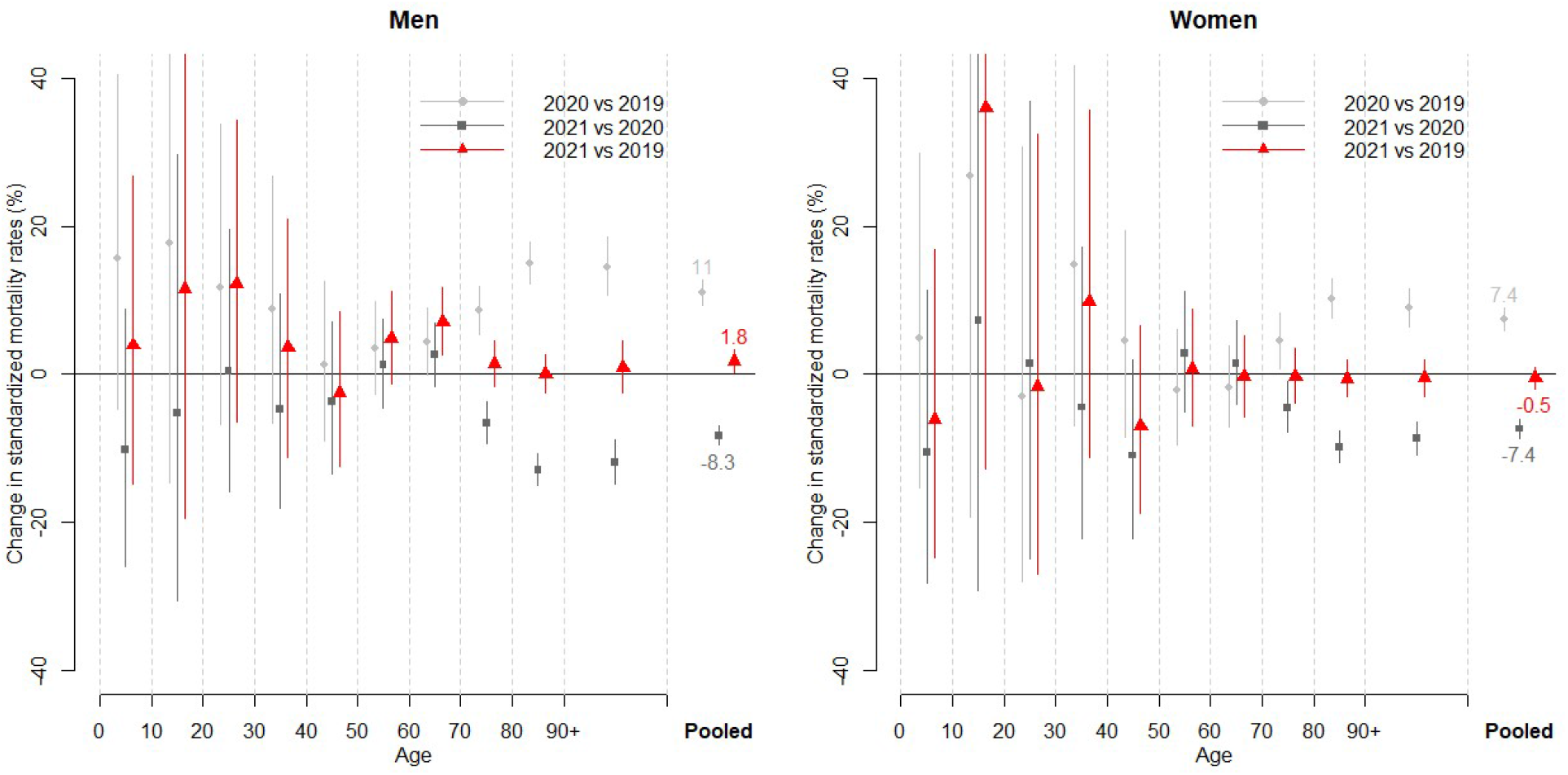
Relative change in standardized mortality rates (reference: January 1^st^ 2021) when comparing 2020 with 2019, and 2021 with 2020 and 2019, in selected age groups, separately for men and women, together with 95% confidence interval (data from the Swiss Federal Statistical Office).

Life expectancy at birth, which reached 81.9 years for men and 85.6 years for women in 2019 (Table 1) after decades of steady increase (about 3 months/year for men and 2 months/year for women), decreased by 10.1 months for men and 5.8 months for women in 2020 reaching 81.0 years for men and 85.1 years for women (Figure 4 and Table 1) [9]. By 2021, this loss has been to a large extent recovered by men, who recovered the 2018 level (life expectancy in 2021 of 81.6 years) and more than recovered by women (life expectancy in 2021 of 85.7 years). We found similar results for life expectancy at age 65 (Fig 4 and Table 1).

**Figure 4.**
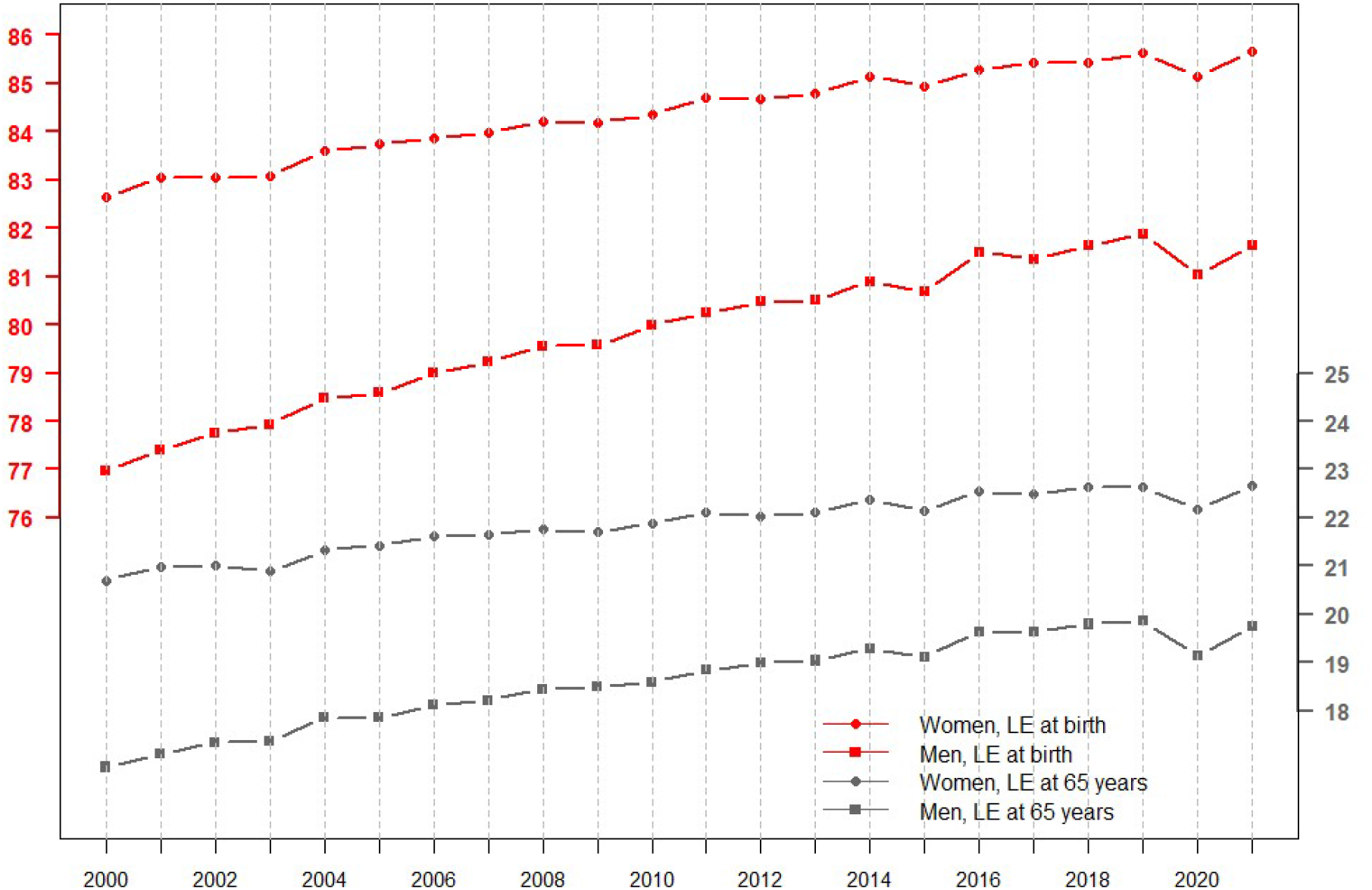
Life expectancy at birth and at 65 years of age in Switzerland for the period 2000-2021 for men and women (data from the Swiss Federal Statistical Office and the Human Mortality Database)

## Discussion

The year 2021 was characterized in Switzerland by a first half of the year with unusually low mortality, due to the absence of any Covid-19 or influenza wave, followed by a second half of the year that was quite standard until an unusual end-autumn wave of mortality, corresponding to the 5th wave of Covid-19. Taken as a whole, 2021 marked an approximate return to 2019 mortality levels. After a 9.2% excess mortality in 2020, standardized mortality rates decreased by 7.8% in 2021, bringing the rate approximately back to the pre-pandemic level. Life expectancy, meanwhile, after declining by 10 months for men and 6 months for women in 2020, recovered in 2021 the 2019 levels for women (85.7 years) while returning to 2018 levels for men (81.6 years). All age groups experienced statistically equivalent mortality in 2020 compared to 2019, with the exception of men aged 60-70 (and to a lesser and not significant extent, men aged 50-60), who still had an excess mortality in 2021 compared to 2019, and who were indeed responsible for the slightly higher mortality observed for men in 2021 compared to 2019.

We also calculated an estimate of excess deaths based on standardized mortality rates. This indicator captures changes in mortality at different ages adjusting for modifications that occur each year in the size and age structure of the population [15-16]. Comparing standardized mortality rates in 2020 and 2021 with those in 2019, we estimated an excess of 6473 deaths in 2020 and an excess of 428 deaths in 2021, leading to an excess of 6901 deaths for the 2020-21 period. Our estimate of the excess death for 2020 was consistent with the excess of 6800 deaths estimated by [4] for Switzerland by applying and projecting a Poisson model on weekly deaths for the years 2016-2019 stratified by age and sex. It was, however, lower than the excess of 8429 deaths estimated by [12] applying and projecting a negative binomial model on weekly deaths of years 2015-2019. Our estimate was also lower than the excess of 8600 deaths estimated via a linear model by [13] for 2020 and the first half of 2021 (considering that the first six months of 2021 had a negative excess of deaths [10,12]). In addition, our estimate of excess death for the entire 2020-21 period was more than two times lower than the excess of 15,500 deaths obtained for Switzerland by a recent analysis of excess mortality for the entire planet [14]. In this paper, the authors estimated the expected mortality for 2020-21 using a global approach over the previous 11 years, weighting six different models, and interpolating the missing information in part of the countries using covariates. We guess that our result is closer to that of [4] because the authors of this study applied their model to age- and sex-stratified deaths, which is roughly equivalent to our standardization. As a complementary analysis, we also estimated the excess deaths for the period 2020-2021 by comparing the standardized mortality rates obtained for these years with those that would be expected based on a continuation of the trend observed in recent years for these rates (Fig 2). However, our results were not the same whether we used ten or twenty years before the pandemic to estimate this trend. Under the first option (linear trend over 2010-19), we estimated an excess of 10377 deaths for the years 2020-21, while the second option (quadratic trend over 2000-19) leaded to an excess of 7903 deaths.

These differences illustrate and confirm the strong dependence of an excess death estimate on the choice of method and time window used to estimate the trends. This is actually the reason why we opted for a factual comparison of mortality after and before the pandemic. For the same reason, it may also be advantageous to quantify the impact of a pandemic on mortality in terms of setbacks along the time axis rather than of projections into the future. Indeed, the setback was the same whether we were looking at standardized rates or life expectancy: both have returned in 2020 to their 2014-16 levels, a five-year setback, while they returned approximately to their 2019 pre-pandemic levels in 2021. Note that a similar phenomenon had been observed for the Spanish flu of 1918. Despite the dramatic loss of life expectancy in 1918 (−10.4 years for men and -8.5 years for women), life expectancy in 1919 (53.7 years for men and 55.9 for women) had largely returned to that of 1917 (54.1 years for men and 57.4 years for women), before the pandemic of Spanish flu.

Regarding the choice of scale to compare mortality across years, we believe that life expectancy has several advantages over excess mortality (or excess deaths). First, being measured in months, a loss of life expectancy is easier to interpret than an excess mortality (in %) or an excess deaths (an absolute number which, on its own, is difficult to apprehend without a good knowledge of the context of a given country). Secondly, by giving more weigh to the (excess) deaths of the young than to those of the elderly, it recognizes that the former are indeed more dramatic than the latter, an essential point when it comes to assess the severity of a pandemic. The conclusions that can be drawn by using one indicator rather than another can be quite different. For example, [12] estimated for Switzerland an excess death about 3 times higher in 1918 (compared to 1917) than in 2020 (compared to 2019). In terms of life expectancy, the ratio between the loss observed in 1918 and that of 2020 is rather 10:1 than 3:1 and, if the two losses are considered in proportion to the (very different) baseline life expectancy of the two periods, the ratio is around 20:1 [8]. The reason for this discrepancy is the different age distribution of the deaths due to the Spanish flu and those caused by Covid-19, the former affecting mostly young people and the latter people over 70. In this regard, it is also interesting to observe that the loss of life expectancy at birth in 2020 (10.1 months for men and 5.8 months for women) corresponds approximately to the loss of life expectancy at age 65 (8.8 months for men and 5.7 months for women). This reflects the fact that almost all the life expectancy loss occurred after age 65.

One limitation of this study is that it uses mortality data that are not yet finalized. However, as we have already noted, the number of deaths reported to date for 2021 is not far from being stabilized, and we believe that, in the situation of a pandemic still in progress, it is important to produce early estimates, even at the risk that they may not be completely accurate. We note, for example, that the estimates we provided one year ago for life expectancy in 2020 [9] proved to be quite close to those obtained when the finalized data became available (with a final discrepancy of only +0.4 months for men and +0.5 months for women) [10]. Thus, although we do not yet have fully definitive results, there is no doubt that mortality in Switzerland in 2021 has largely returned to its pre-pandemic level. An analysis of the possible reasons responsible for this favorable trend, including vaccination, more experience in treating the disease, improved patient management, harvesting effects, and the emergence of less virulent variants of the virus, is beyond the scope of this study.

## Data Availability

Swiss FSO:
DEATHS:
https://www.bfs.admin.ch/bfs/fr/home/statistiques/population/naissances-deces/deces.html
POPULATION:
https://www.bfs.admin.ch/bfs/fr/home/statistiques/population/effectif-evolution/age-etat-civil-nationalite.assetdetail.18344199.html
Human Mortality Database (HMD):
https://www.mortality.org/

## References

1. Beaney T, Clarke J, Jain V, et al. (2020). Excess mortality: the gold standard in measuring the impact of COVID-19 worldwide? Journal of the Royal Society of Medicine, 113: 329–334

2. Woolf SH, Chapman DA, Sabo RT, Zimmerman EB. Excess Deaths From COVID-19 and Other Causes in the US, March 1, 2020, to January 2, 2021 (2021). JAMA, 325:1786–1789. doi:10.1001/jama.2021.5199

3. Ahmad F, Cisewski J, Minino A, Anderson R (2021). Provisional mortality data – United States, 2020. Morbidity and Mortality Weekly Report, 70: 519–522

4. Islam N, Shkolnikov VM, Acosta RJ et al. (2021) Excess deaths associated with covid-19 pandemic in 2020: age and sex disaggregated time series analysis in 289 high income countries. BMJ 373: n1137 http://dx.doi.org/10.1136/bmj.n1137

5. Aburto JM, Kashyap R, Schöley J, et al. (2021) Estimating the burden of the COVID-19 pandemic on mortality, life expectancy and lifespan inequality in England and Wales: a population-level analysis. J Epidemiol Community Health, 75: 735–740. doi: 10.1136/jech-2020-215505

6. Andrasfay T, Goldman N. (2021) Reductions in 2020 US life expectancy due to COVID-19 and the disproportionate impact on the Black and Latino populations. Proc Natl Acad Sci USA, 2: 118:e2014746118. doi: 10.1073/pnas.2014746118

7. Heuveline P, Tzen M (2021). Beyond deaths per capita: comparative COVID-19 mortality indicators. BMJ Open, 11:e042934. doi:10.1136/bmjopen-2020-042934.

8. Rousson V, Paccaud F, Locatelli I (2022) A comparison of the losses of life expectancy at birth between the 2020 and 1918 pandemics in six European countries. Vienna Yearbook Population Research. Submitted

9. Locatelli I & Rousson V (2021). A first analysis of excess mortality in Switzerland in 2020. Plos One. https://doi.org/10.1371/journal.pone.0253505

10. Locatelli I, Rousson V. Mortality in Switzerland 2020-2021 (2021). Center for Primary Care and Public Health (Unisanté), University of Lausanne, Lausanne (Raisons de Santé: Les Essentiels 35) http://dx.doi.org/10.16908/rds-essentiels/

11. Aburto J, Schöley J, Kashnitzky I, et al. (2021). Quantifying impacts of the COVID-19 pandemic through life expectancy losses: a population-level study of 29 countries. International Journal of Epidemiology, 51: 63– 74, https://doi.org/10.1093/ije/dyab207

12. Staub K, Panczak R, Matthes KL, et al. (2022) Historically High Excess Mortality During the COVID-19 Pandemic in Switzerland, Sweden, and Spain. Ann Intern Med, 1:M21–3824. doi: 10.7326/M21-3824

13. Karlinsky A and Kobak D (2021) Tracking excess mortality across countries during the COVID-19 pandemic with the World Mortality Dataset. eLife; 10:e69336 DOI: 10.7554/eLife.69336

14. COVID-19 Excess Mortality Collaborators (2022) Estimating excess mortality due to the COVID-19 pandemic: a systematic analysis of COVID-19-related mortality, 2020-21. THE LANCET DOI:https://doi.org/10.1016/S0140-6736(21)02796-3

15. Curtin L, Klein R (1995). Direct standardization (age-adjusted death rates). Healthy People 2000 Statistical Notes, 6, 1–10.

16. Fleiss JL, Levin B, Cho Paik M (2003). Statistical methods for rates and proportions. Wiley Series in Probability and Statistics Eds.

17. Friedman M (1982). Piecewise exponential models for survival data with covariates. Annals of Statistics, 10, 101–113.

18. Preston Samuel H.; Patrick Heuveline; Michel Guillot (2001). Demography: measuring and modeling population processes. Blackwell Publishers. ISBN 1-55786-214-1.

